# Underreporting of death by COVID-19 in Brazil’s second most populous state

**DOI:** 10.1101/2020.05.20.20108415

**Authors:** Thiago Henrique Evangelista Alves, Tafarel Andrade de Souza, Samyla de Almeida Silva, Nayani Alves Ramos, Stefan Vilges de Oliveira

## Abstract

The COVID-19 pandemic brings to light the reality of the Brazilian health system. The underreporting of COVID-19 deaths in the state of Minas Gerais (MG), where is concentrated the second largest population of the country, reveals government unpreparedness, as there is a low capacity of testing in the population, which prevents the real understanding of the general panorama of Sars-Cov-2 dissemination. The goals of this research are to analyze the causes of deaths in the different Brazilian government databases (ARPEN and SINAN) and to assess whether there are sub-records shown by the unexpected increase in the frequency of deaths from causes clinically similar to COVID-19. A descriptive and quantitative analysis of the number of COVID-19 deaths and similar causes was made in different databases. Ours results demonstrate that the different official sources had a discrepancy of 209.23% between these data referring to the same period. There was also a 648.61% increase in SARS deaths in 2020, when compared to the average of previous years. Finally, it was shown that there was an increase in the rate of pneumonia and respiratory insufficiency (RI) by 5.36% and 5.72%, respectively. In conclusion, there is an underreporting of COVID-19 deaths in MG due to the unexplained excess of SARS deaths, Respiratory insufficiency and pneumonia compared to previous years.

## Introduction

Coronavirus 2 (COV-2) is a new betacoronavirus related to Severe Acute Respiratory Syndrome (SARS) that emerged in December 2019 in China and became a pandemic in March 2020 due to its high infection and mortality rates[1] [2] [3].

COVID-19 was the official name given by the World Health Organization (WHO) to the disease caused by the new coronavirus of 2019(SARS-CoV-2) [1].

The first epicenter of COVID-19 was observed in Wuhan, the capital of Hubei, China, in December 2019 based on the several pneumonia cases notifications [4].

Since then, COVID-19 has rapidly spread around the world and, as of May 12th 2020, more than 4.4 million cases of the disease have been confirmed, causing over 299,000 deaths worldwide. [5]. Of this total, Brazil has reported more than 188,000 cases and over 13,000 deaths, according of Coronavírus Brasil database [6].

COVID-19 is classified according to the symptoms’ severity. Patients with the mild form (80% of the cases) present fever, dry cough, chills, malaise, muscle pain,and sore throat. Patients with moderate form present fever, respiratory symptoms, and radiographic characteristics. Severe patients (5% of the cases) manifest dyspnea (> 30 bpm), low oxygen saturation (<93%) and low PaO2/FiO2 ratio (< 300 mmHg), and may evolve to a respiratory failure, septic shock, and multiple organ failure [7] [8] [9].

Furthermore, increased age and the presence of comorbidities, such as hypertension, diabetes, and coronary disease, are associated with mortality in COVID-19 patients [10] [11].

The accurate diagnosis of COVID-19 is carried out by searching the genetic material of the virus and, in a complementary way, by imaging methods. Computed tomography and radiographs can identify lesions in the lungs due to viral multiplication [12] [13].

Laboratory confirmation is essential for the timely management of cases to avoid the spread of transmission. However, Brazil is far below the ideal number of tests for COVID-19, as there are not enough laboratory inputs to understand the overall panorama of the virus’ spread. Furthermore, confirmatory molecular tests depend on the availability of imported reagents,whichare globally scarce, and on government investments that prioritize this strategy. This scenario leads Brazil to have a delay in the number of COVID-19 cases and deaths confirmations. These aspects become more aggravated when the patient evolves to death, because the effectiveness of tests, for these cases, is even more difficult. In addition, the recommendation is to collect blood and sputum to perform the culture, since these samples have a higher viral load – considering the studies done to date [14].

The difficulty regarding death registration have been also presented in the state of Minas Gerais, which, by the end of April 2020, had 584 suspected deaths notifications, of which 81 (13%) had not yet been confirmed or discarded [15]. Thus, it is possible to state that there is a disparity between the real number of COVID-19 deaths and the numbers that are reported in different Brazilian sources of information, since not all deaths have been tested for confirmation or exclusion and are potentially being confirmed by others causes than COVID-19.

The present study aims to analyze the death causes in the notary records and in the Brazilian National disease notification system records, and thus evaluate the sub-registries and the possible increase in the frequency of deaths with clinically compatible causes to COVID-19 in the Minas Gerais territory.

## Method

This study is a descriptive and quantitative analysis of the deaths records clinically compatible with COVID-19, registered in the notary offices records and in the Brazilian National Disease Notification System Records (SINAN) of the state of Minas Gerais (MG), Brazil.

The state of MG has an estimated population of 21,168,791 people in a territory of 586,521.121 km^2^, having the second largest population and being the fourth largest state in the country [16]. Its Human Development Index (HDI) is 0.731, with a population composed of approximately 22.25% from 0 to 15 years old, 69.31% from 15 to 64 years old, and 8.12% over 65 years old [17]. For this study, the notary offices records were analyzed from January to April of 2020 in the state of MG. Additionally, to assess the deaths excess in this period according to their causes, information from the SINAN was accessed referring to the range of years 2017 to 2019.

The notary data were obtained from the Civil Registry Transparency Portal, which is a free access platform developed to provide information about births, marriages, and deaths. Due to the COVID-19 pandemic, these data are being grouped in the Special sections COVID-19 and the COVID Registral Panel made available on ARPEN database [18]. The information presented here (accessed on 05/05/2020) is based on Death Certificates (DD), presenting only one cause for each death certificate [19].

To evaluate sub-registrations in the different Information Systems in Brazil, SINAN data were collected through the InfoGripe platform of the Oswaldo Cruz Foundation (Fiocruz) InfoGripe database [20] is an initiative that aims to monitor and present alert levels for reported cases of Severe Acute Respiratory Syndrome (SARS) in SINAN [20]. The data in this system were compared with the notary data. On this platform, the records of SARS and COVID-19 were selected on 05/07/2020 according to the Epidemiological Week (EPI Week) from 1 to 18 of the years 2017 to 2020 for the state of Minas Gerais.

We also evaluated the death excess from causes that present clinical compatibility with COVID-19, according to the following etiology: Severe Acute Respiratory Syndrome (SARS), Pneumonia, Respiratory insufficiency (RI), Sepsis (sepsis/septic shock), Indeterminate Causes (deaths related to respiratory diseases, but not conclusive), and Other deaths (all other types of deaths that are not listed above) [19].

The data were collected and analyzed in spreadsheet by descriptive statistics and presented in raw numbers, relative frequency, and central tendency measures. To assess the death excess per EPI week, the average, minimum and maximum values of deaths from the years 2017 to 2019 were calculated and confronted with diseases that presented changes in the pattern of distribution in the fourth quarter of 2020. All graphs were prepared using GraphPadPrism 7 software (GraphPad Software, Inc. San Diego, CA).

## Results

A total of 201 COVID-19 deaths were identified in the notary records and this number differs in 209.23% of the deaths registered in the SINAN at the same period (Table 1).

**Table 1:** Number of COVID-19 deaths according to the information in the notary offices records and the Brazilian National Disease Notification System Records (SINAN).

| COVID-19 | January |  | February |  | March |  | April |  |
| --- | --- | --- | --- | --- | --- | --- | --- | --- |
|  | N | % | N | % | N | % | N | % |
| Notary offices records | 0 | 0 | 0 | 0 | 23 | 11.44 | 178 | 88.56 |
| SINAN records | 0 | 0 | 0 | 0 | 23 | 35.38 | 42 | 64.62 |

The evaluation of the death causes on the notaries’ offices showed an increase in the frequency of SARS deaths in 2020 in relation to the number of deaths from the same disease in 2019. There was also a slight increase in the number of pneumonia and respiratory failure deaths in 2020 between January and March (Figure 1).

**Figure 1:**
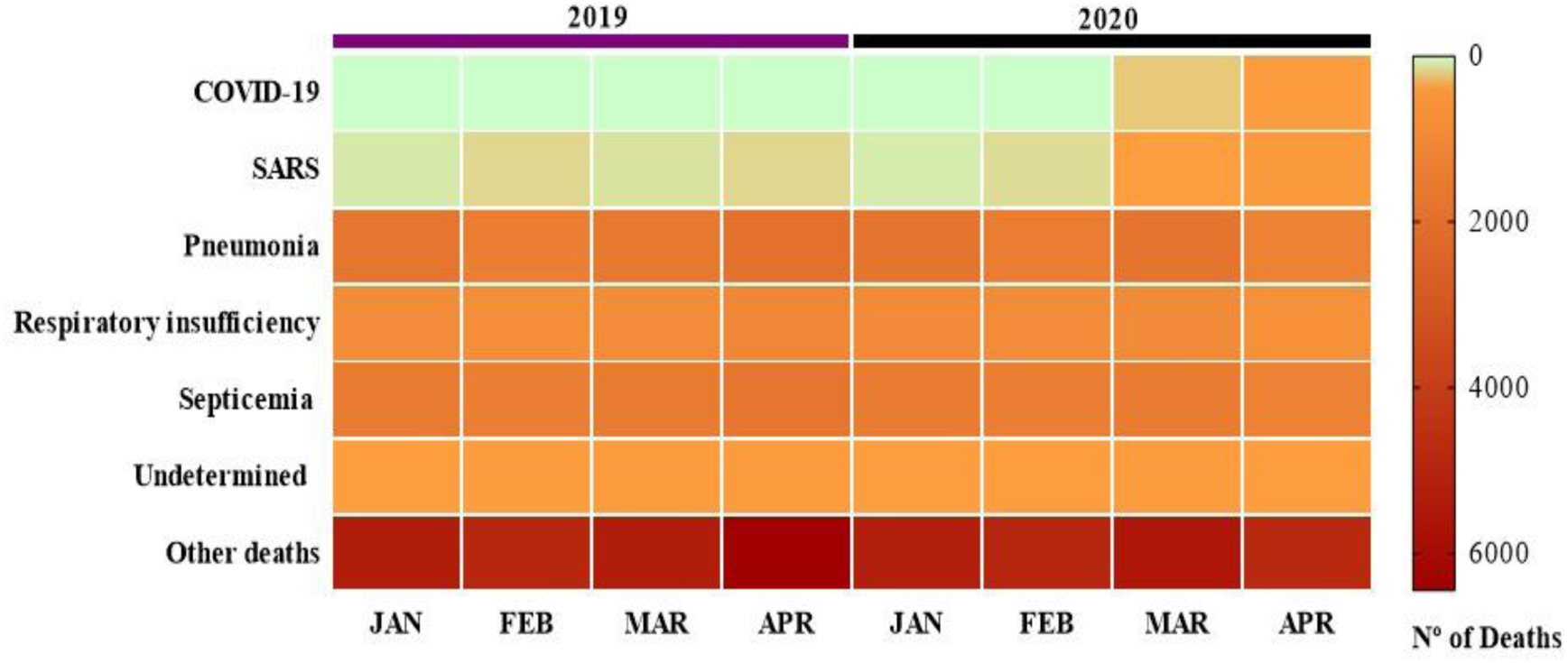
Distribution of deaths from COVID-19, Severe Acute Respiratory Syndrome (SARS), Pneumonia, Respiratory insufficiency (RI), Sepsis (sepsis/septic shock), Indeterminate Causes (deaths related to respiratory diseases, but not conclusive), Other Deaths (all other types of deaths that are not listed above), according to the notary offices records from January to April 2019 and 2020, in the state of Minas Gerais, Brazil.

The increase of SARS case sin 2020 was in the order of 152.7% compared to 2019. Regarding to the elevation in the rates of pneumonia and respiratory insufficiency from January to March compared to the same period in 2019, it was around 5.36% and 5.72%, respectively.

As shown in Figure 2, when analyzing the excess of deaths in 2020 according to epidemiological weeks 1st to 18^th^, there was an increase of 648.61% in SARS deaths compared to the average of previous years (2017/2019). Such ascendancy of SARS deaths was observed from epidemiological week number 10 and the records of COVID-19 deaths in the state of MG are reported from epidemiological week number 12.

**Figure 2:**
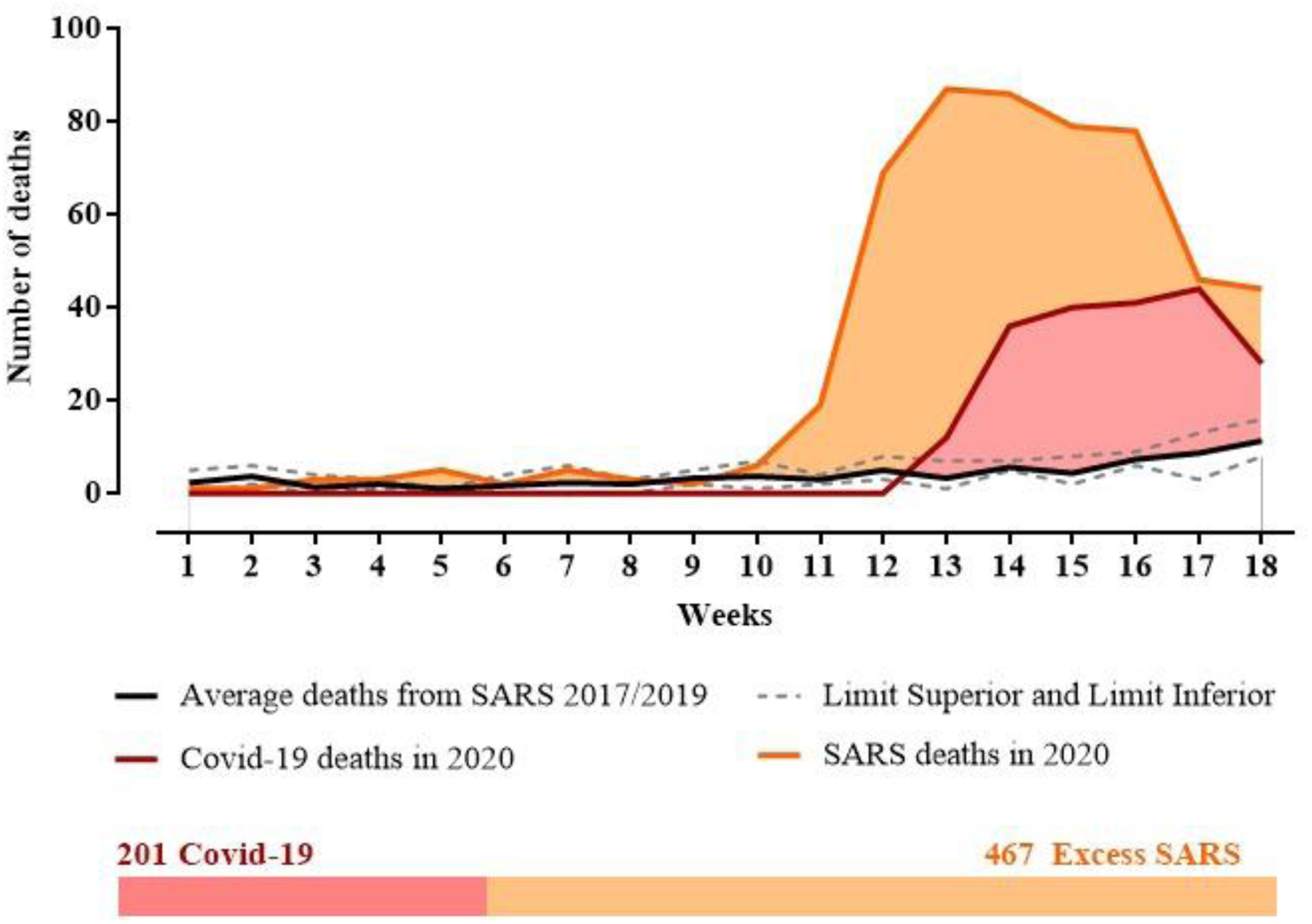
Evaluation of COVID-19 deaths from ARPEN and excess deaths from SARS in 2020, during epidemiological weeks 1st to 18^th^, comparing the data from the SINAN (2017 to 2019) in the state of Minas Gerais, Brazil.

## Discussion

The study points out divergences of information between different death registrations systems. The important increase in SARS deaths that started earlier than those from COVID-19, in epidemiological week 10, is also highlighted, suggesting the underreporting of COVID-19 deaths in the state of Minas Gerais.

The COVID-19 situation is particularly challenging because, besides being a new and unprecedented disease, it is also capable of triggering other conditions, such as pneumonia and SARS, which can be characterized as the main cause of death. In other words, the COVID-19 may be the underlying cause, that is, it may not be the direct cause of death that has been registered. In this perspective, there is a subjectivity bias, since the physician can attest or not the death from COVID-19 according to his clinical knowledge without the need of laboratory tests [21]. This finding corroborates with data from Hubei, China and Northern Italy, where mortality calculations were adjusted for the biases of preferential verification, symptomatic and severe cases, and delay in death records. An increase in the mortality rate was found, which confirms the existence of underreporting COVID-19 deaths in those regions [22].

In relation to underreporting in Brazil, the Ministry of Health (MH) reports that the number of under-reported deaths is low according to the Mortality Information System (MIS), because states and municipalities are advised to include deaths from COVID-19, either confirmed cases or only suspects, in the system as a priority, in order to advance analysis of these cases [23]. However, our results show that there is a significant underreporting of the occurrences by COVID-19, given the excess of SARS deaths.

Another issue that should be analyzed is that although the Civil Registry Information Center takes into consideration both confirmed deaths and suspects, the MH discloses in its reports only the laboratory proven COVID-19 deaths [23]. However, suspect deaths need to be considered in the count, even though it is noted that they have not been confirmed. This is stated because it is known that many of these deaths will not be able to be analyzed, given the difficulties in collecting, transporting, and wrapping the *post-mortem* samples. Thus, if they are not mentioned, there will be a relaxation of the real situation in Brazil and, consequently, in the state of MG.

The Brazilian MH also points out that in the same death certificate more than one cause of death can be described, so that the record of COVID-19 can be associated with other diseases. However, the Civil Registry Transparency Portal presents these causes separately, even those included or registered in the same death certificate. Thus, one cannot only add up the deaths made available on the portal by the different diseases, because they would generate false over-notification. A thorough investigation must be made when considering each death and the causes that were cited in the death certificate [23]. However, according to the hierarchical criteria exposed in the Civil Registry Transparency Portal, only one cause of death is selected to make the count, and not all the causes present in the same death certificate [24], which validates the data exposed in this platform and the information presented here.

It is worth noting that the different systems of deaths registration of the government, such as the municipalities and states, are not fully connected and that several of them depend on manual labor to be registered. This is capable of causing discrepancies and delays in data traffic and, consequently, in the production of timely and reliable information.

WHO has been advising countries on the need to expand laboratory testing capacity as a strategy to overcome the pandemic [25]. This action will enable the real knowledge of a population’s immunity, providing reliable statistics for a better understanding of the circulation of the disease. Consequently, strategies to control the pandemic and even the relaxation of non-pharmacological measures, such as social isolation and quarantines, may be proposed.

In Brazil, a network formed by referenced laboratories was established to help fight COVID-19 [14]. However, the country is far below the optimal number of tests for COVID-19, as there are not enough tests to have a reliable panorama of the real number of cases and deaths. This scenario leads Brazil to have a delay in accounting the records of COVID-19.

In conclusion, our results reveal that COVID-19 deaths in the state of Minas Gerais are higher than the official statistics presented. In view of these aspects, it is necessary to expand Brazil’s diagnostic capacity, which will allow us to recognize the real number of COVID-19 deaths and cases in Minas Gerais.

## Data Availability

All data used in these articles are publicly accessible and can be consulted

## Acknowledgement

Thanks to Gabriela Geraldo Mendes and Adélio Tiago da Mota for the collaborations. Thanks to the Department of Collective Health of the Faculty of Medicine of the Federal University of Uberlândia for the encouragement.

## Notes

### Competing Interest Statement

The authors have declared no competing interest.

### Funding Statement

This study did not have funding.

